# Nurses’ Lived Experiences of Caring for Patients During the 2022 Ebola Virus Disease Outbreak in Uganda: A Qualitative Study

**DOI:** 10.64898/2026.08.25.26361289

**Authors:** Gift Mushabe, Patience Muwanguzi, Sarah Joselyn Nalubega, Bridget Atuhaire, Maureen Namugaya, Racheal Nabunya

**Affiliations:** School of Health Sciences, College of Health Sciences Makerere University, P. O. Box 7072, Kampala, Uganda; Humanitarian and Conflict Response Institute, University of Manchester, Manchester, UK; African Centre for Health Equity Research & Innovation, P.O. Box 5259, Kampala; School of Medicine, College of Health Sciences Makerere University, P.O Box 7072, Kampala, Uganda

**Keywords:** Ebola Virus Disease, Lived Experiences, Nursing, Uganda, Qualitative Research

## Abstract

**Background:** Nurses play a pivotal role during infectious disease outbreaks, including Ebola Virus Disease (EVD), often placing themselves at significant personal and professional risk. While nurses are central to Ebola response efforts, their lived experiences, particularly in Uganda, remain underexplored. This study aimed to explore the lived experiences of nurses who provided care to EVD patients during the 2022 outbreak at Mubende Regional Referral Hospital.

**Methods:** An exploratory qualitative design was employed using in-depth interviews with seven nurses who worked in the Ebola Treatment Unit (ETU). Participants were selected through purposive and snowball sampling. Data were analysed thematically using a phenomenological approach to capture the emotional, psychological, and professional dimensions of nurses’ experiences.

**Results:** Findings revealed a complex interplay of positive and negative experiences. Positive experiences included joy from patient recovery, strong teamwork, professional growth, and enhanced training in infection prevention and control (IPC). Nurses reported increased confidence, new skills, and opportunities for career advancement, including recognition from international agencies.

Conversely, negative experiences dominated the narratives. These included severe resource constraints, overwhelming workload, physical exhaustion, and psychological distress, particularly from witnessing patient suffering and death. Participants described post-traumatic stress symptoms and constant fear of infection and death, exacerbated by inadequate early-stage preparation. Social isolation and disrupted family life were also common, with some nurses concealing their work to avoid distressing loved ones or experiencing stigma within their communities. Additional challenges included team tensions, absenteeism, and disparities in compensation and recognition between existing staff and external recruits.

**Conclusion:** The study highlights the profound emotional and professional toll on nurses during the 2022 Ebola outbreak in Uganda. While moments of resilience and growth were evident, structural and systemic challenges significantly impacted nurses’ well-being and care delivery. To better support frontline health workers, future outbreak responses must prioritize fair compensation, psychosocial support, consistent training, and improved resource allocation. Insights from this study can inform policies aimed at strengthening workforce readiness and retention during health emergencies.

## Background

Caring for patients with Ebola virus disease (EVD) requires specialized healthcare skills and meticulous management namely; screening and triage, isolation precautions, patient handling, and infection prevention and control measures (1, 2), due to the disease’s highly contagious nature and severe outcomes (3). Ebola virus disease, formerly known as Ebola haemorrhagic fever (EHF), is a highly lethal infection affecting both humans and non-human primates(4). The global case fatality rate (CFR) for EVD is estimated at 24.5% (95% CI 0.0–67.9%) (5). However, in sub- Saharan Africa (SSA), the CFR is significantly higher, at 61.3% (95% CI 52.8– 69.6%), reflecting the region’s heightened vulnerability to the disease (5). A total of 35 EVD outbreaks have been reported in SSA amounting to 164,894 cases between 1976 and 2022 (5). The Democratic Republic of Congo has the most outbreaks, followed by Uganda with six (6). The most destructive outbreak in Uganda occurred in 2000 in Gulu district, with 425 cases and 224 deaths, 22 of which were healthcare workers. Subsequent outbreaks, such as the 2007 Bundibugyo incident, infected 14 healthcare workers, and the most recent outbreak in September 2022 in Mubende district led to 164 cases and 77 deaths, with 19 healthcare workers infected and 7 fatalities among them (7, 8).

Healthcare professionals play a critical role in providing care to EVD patients during EVD outbreaks. Nurses, who make up the majority of the health workforce (59% globally) (9), are at the forefront of the healthcare system’s response to epidemics and pandemics(10). They are directly involved in patient care, including assessment, specimen collection, monitoring response to treatment, and assisting with activities of daily living (11). This puts them at a high risk of exposure to this viral disease and of dying as a result (12). Despite their professional obligation to care for the community during a pandemic or epidemic, many nurses are concerned about the risk of infection, potential virus transmission to family members, stigma associated with their roles, and personal freedom restrictions.(13).

Nurses caring for EVD patients have reported both positive and negative experiences. Positive experiences may include the joy of seeing patients recover, working with a supportive team and positive environment, and fulfilling their professional duties (14). However, these are often overshadowed by negative experiences, such as resource limitations, lack of well-defined roles, and mental health challenges like stress, depression, anxiety, post-traumatic stress disorder (PTSD), and burnout. These challenges affect their concentration, comprehension, decision-making abilities, and willingness to provide care, potentially affecting the overall effectiveness of infectious disease management (15, 16). Understanding these experiences is crucial to ensuring that nurses are well supported, enabling them to remain in the workforce and continue providing high-quality care during times of heightened community health needs (13).

While some studies have explored the experiences of healthcare workers during EVD outbreaks in Uganda, for example, a study done in 2019 (17) and in 2022(8), there remains a significant gap in the literature regarding the lived experiences of nurses. Most studies have focused on the epidemiology of EVD(4), its emergency state in Uganda(18), or the experiences of survivors(19, 20), leaving the lived experiences of frontline nurses underexplored.

Given the crucial role nurses play in managing Ebola outbreaks, it is essential to understand their personal and professional challenges while on the frontlines. This study, therefore, explored the lived experiences of nurses caring for patients with EVD at Mubende Regional Referral Hospital. By shedding light on these experiences, this research sought to provide insights that could enhance support systems, improve nursing practices, and ultimately lead to better healthcare outcomes during future outbreaks.

## Methods

### Study design and setting

A qualitative phenomenological study was conducted in April 2023 to explore the lived experiences of nurses providing care to EVD patients at Mubende Regional Referral Hospital following the outbreak in 2022. This hospital is a government-owned facility located in the central region of Uganda that serves as the primary referral centre for several districts. During the Ebola outbreak, Mubende Regional Hospital played a crucial role as a treatment centre for suspected EVD cases for isolation and treatment. It established an Ebola Treatment Unit (ETU) to provide specialized care for patients affected by the virus(21). It comprised 48 beds, with 24 allocated for suspected cases and 24 for confirmed cases(22). Therefore, Mubende Regional Hospital was chosen due to its direct involvement in the care of EVD patients. Nurses were eligible to participate in the study if they were involved in the care of patients with Ebola virus disease. Approval to conduct the study was sought from the Makerere University School of Health Sciences Research Ethics Committee (Ref. number: MakSHSREC-2023-494) and the Mubende Hospital Research and Ethics Committee. With permission, the team gained access to the study participants and the hospital.

### Data collection

We applied the Snowball sampling technique, also known as chain referral sampling. This sampling method was appropriate for hard-to-reach populations, such as nurses who were involved in the care of Ebola virus disease patients. The initial participants were identified through purposive sampling, and then these were asked to refer other potential participants who met the study’s inclusion criteria. This process continued until the desired sample size was reached.

Data was collected through in-depth interviews using an interview guide (Supplementary file 1). The guide comprised open-ended questions that allowed participants to provide detailed descriptions of their experiences. This guide was developed by the principal investigator based on the literature review and expert consultations in the field of Ebola care. It underwent pilot testing to ensure comprehensive coverage and appropriate interview length and flow.

Participants were briefed on the study’s objectives and assured of confidentiality before providing consent for participation. The interviews were conducted face-to-face by the researcher in a private location convenient for the participants The interviews lasted between 30 and 60 minutes and were audio-recorded with participants’ consent using an audio recorder. We determined the sample size based on data saturation principle, and we ceased recruitment when no new information emerged from participant interactions.

### Data analysis

The audio files and the transcripts were securely stored under lock and key on a password protected computer and only accessible to the research team. Each participant was assigned a unique identification number to protect their privacy. The audios were transcribed verbatim. The research team read and re-read the transcripts to familiarise and ensure the accuracy and proper interpretation of the transcripts Data was manually analysed using thematic analysis, where phrases and sentences related to nurses’ lived experiences of caring for patients with EVD were coded in the margin of the transcript sheet. The codes were thoroughly reviewed, and those with similar contents were grouped into subthemes, and then into themes.

## Results

### Socio-demographic characteristics of the participants

Seven (7) nurses were interviewed in the study. Of these, 3 (42.9%) were aged between 30 and 40, and the majority, 5 (71.4%), had more than 5 years of working experience. The highest nursing qualification attained was a bachelor’s degree. Six (85.7%) had cared for Ebola patients for less than 2 months. (Table 1)

**Table 1:**
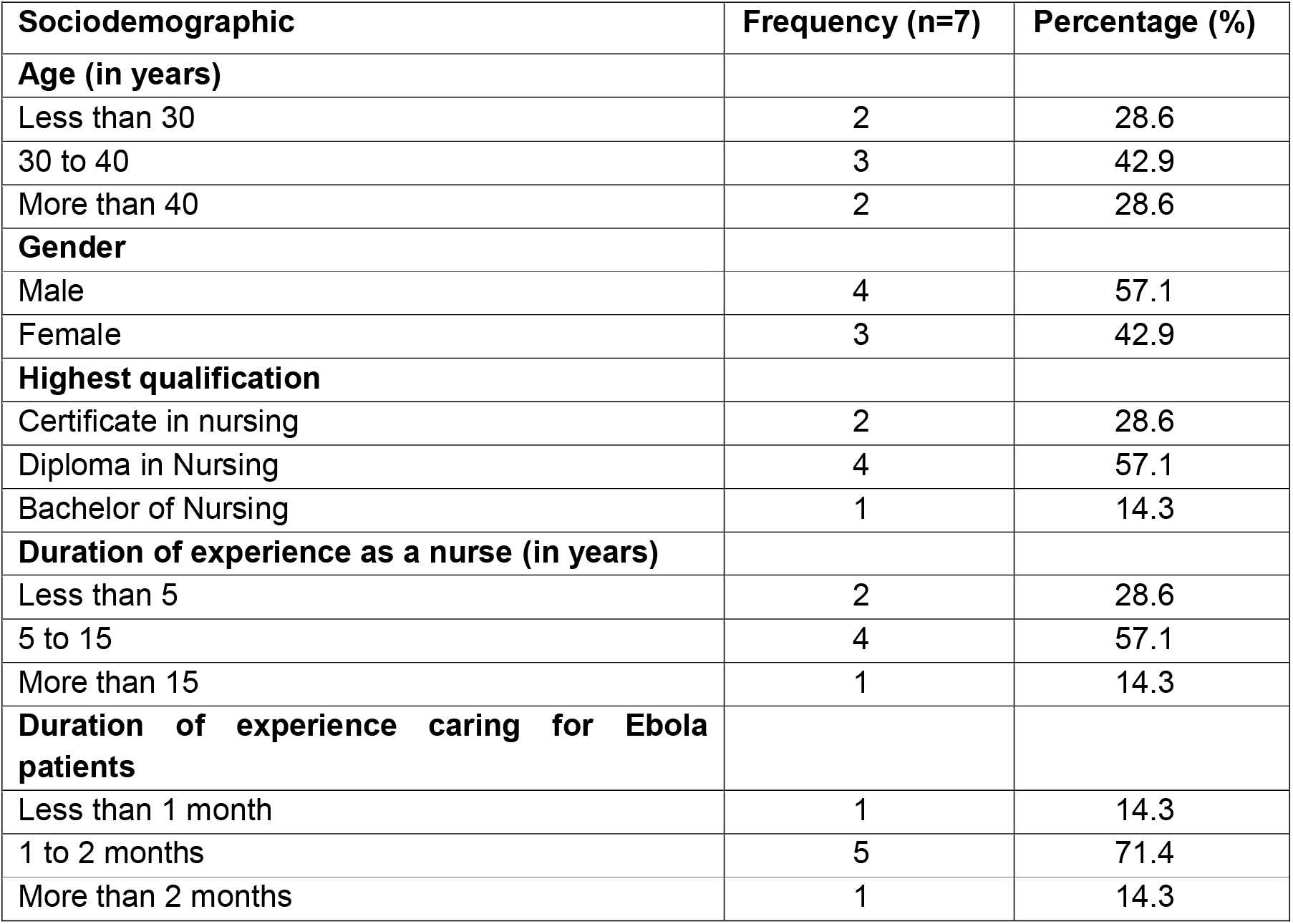
Socio-demographic characteristics of nurses caring for patients with Ebola virus disease in Mubende Regional Referral Hospital n=7

| Sociodemographic | Frequency (n=7) | Percentage (%) |
| --- | --- | --- |
| **Age (in years)** |  |  |
| Less than 30 | 2 | 28.6 |
| 30 to 40 | 3 | 42.9 |
| More than 40 | 2 | 28.6 |
| **Gender** |  |  |
| Male | 4 | 57.1 |
| Female | 3 | 42.9 |
| **Highest qualification** |  |  |
| Certificate in nursing | 2 | 28.6 |
| Diploma in Nursing | 4 | 57.1 |
| Bachelor of Nursing | 1 | 14.3 |
| **Duration of experience as a nurse (in years)** |  |  |
| Less than 5 | 2 | 28.6 |
| 5 to 15 | 4 | 57.1 |
| More than 15 | 1 | 14.3 |
| **Duration of experience caring for Ebola patients** |  |  |
| Less than 1 month | 1 | 14.3 |
| 1 to 2 months | 5 | 71.4 |
| More than 2 months | 1 | 14.3 |

### Lived experiences of nurses caring for patients with Ebola virus disease

Two themes emerged, that is positive and negative experiences. A summary of the themes, subthemes, and categories is shown in Figure 1.

**Figure 1:**
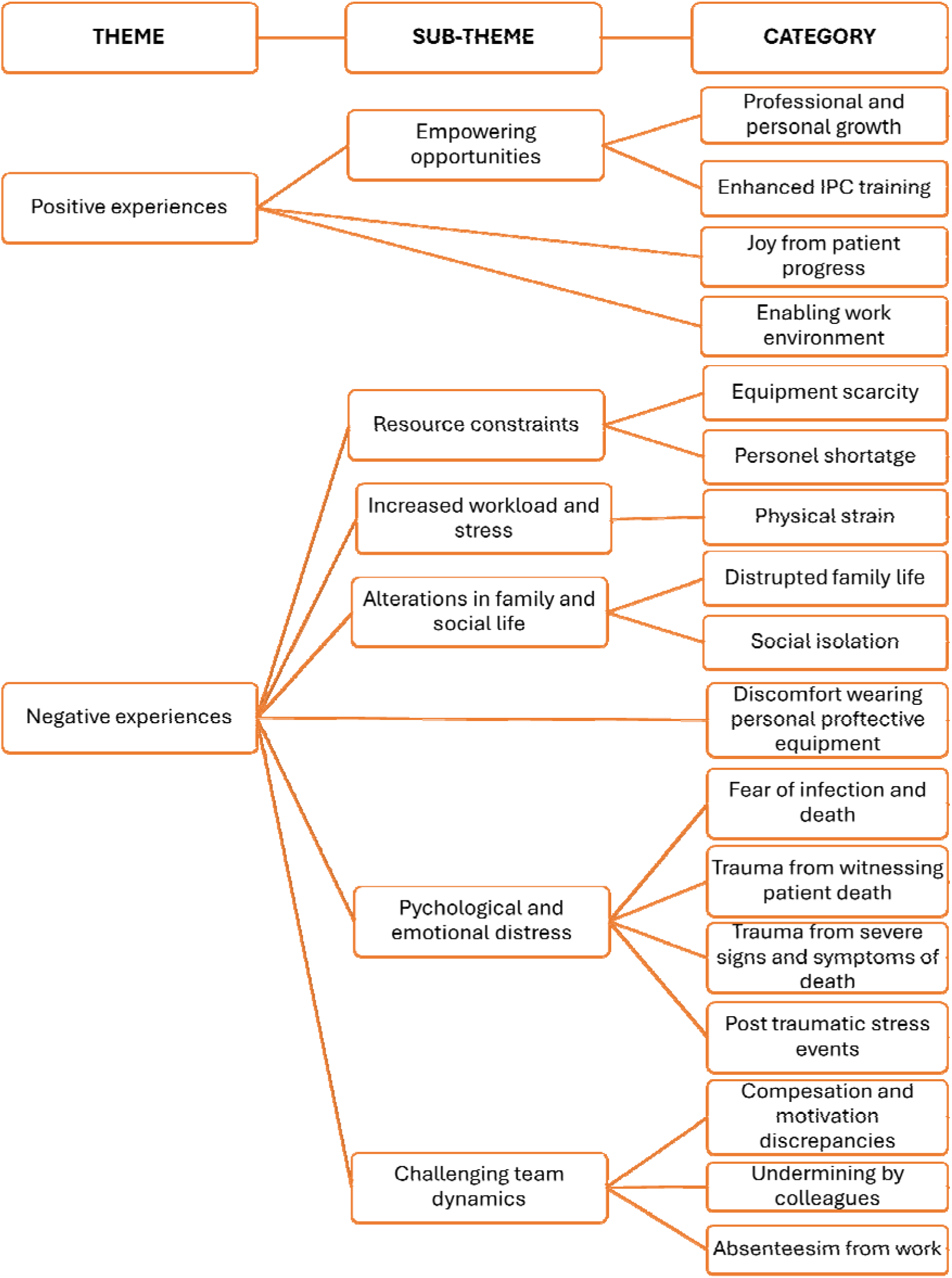
A summary of the subthemes and categories for the lived experiences of nurses caring for Ebola patients

### Positive experiences

One subtheme emerged from the analysis: empowering opportunities. This subtheme comprised two categories: professional and personal growth, and enhanced IPC training. Additionally, related categories under this theme were joy from patient progress and an enabling work environment.

#### Joy from patient progress

The nurses reported feelings of happiness and satisfaction when patients showed improvement or fully recovered from Ebola. They described these moments as their best days in the ETU, particularly when patients in critical condition defied the odds and recovered.

> "My best day in the ETU—actually, my best days—were during those first three weeks. Most of the patients we received were in bad shape: they were weak, they were wet, but we managed to help them recover and discharge them. That brought us so much happiness—the whole team was joyful." - Participant 03

Another participant reported that the greatest joy was when they discharged all the patients, and the ETU remained empty

> *"There was one mother left in the ETU. When we discharged her, she was full of joy—and so were we. We danced and celebrated because the ETU was finally empty. It was such a joyful moment. You know, it’s incredibly uplifting to discharge a survivor of Ebola, especially knowing how deadly the disease is."* -Participant 01

#### Enabling work environment

Participants highlighted the importance of having a committed and active team. They watched and protected one another from possible exposure to Ebola virus disease and this helped most of them not to acquire the disease throughout the whole period of providing care. They further stated that this support from colleagues was crucial in creating a safe and efficient work environment.

> *"For me, the first three weeks were actually very good. The team we had was committed and very focused. All of us were active—we knew exactly what we were managing. Even though resources were limited, the teamwork made everything easier. We supported one another—no one acted like they were always right or wrong. If you saw a colleague struggling, you helped them. That sense of unity and protecting each other is what really helped us through."* -Participant 03.

One participant reported that the team supported him especially when he grew emotional and provided care beyond the standard protocols because the patient, he was taking care of, was his friend.

> *"I became a bit emotional—I wanted to spend more time with the patient. I wanted to clean him up, feed him, and collect some samples that hadn’t yet been taken, but time had run out. They noticed I was taking it personally and going beyond the standard protocols, even staying past my shift. Eventually, they had to pull me out and force me to stop. That reduced my risk of exposure."* -Participant 04.

### Empowering opportunities

#### Professional and personal growth

Some participants reported that, because of their work, they have had opportunities for professional growth, including being added to international databases. As a result, they are now eligible for future deployments in the event of outbreaks in other countries. Others shared the opportunities they gained and the recognition they received during their work. Many of them reported their social networks increased while others reported collaborating with personnel from outside Uganda and signing contracts with big organizations.

> *"Even now, my name is in the World Health Organization database. If there’s another Ebola outbreak—say in Sierra Leone—they can call me to help. I’ll be more confident because I learned a lot during that outbreak. I gained knowledge, not just in patient care, but also in research."* -Participant 1

> *"I didn’t remain the same in terms of skills—and even my knowledge grew. Through working and interacting with people from different places, including those at the international level, I also expanded my social networks."* - Participant 05

#### Enhanced IPC training

The training provided by the Ministry of Health and WHO helped the nurses feel more prepared and confident in managing the outbreak. Most of them reported that this training was both practical and theoretical, and it was continuous throughout the period of caring for patients. The training helped to improve their knowledge and confidence in infection prevention and control (IPC) and the care provided during infectious diseases. It also prepared the nurses for any Ebola outbreak, reduced the risk of transmission, and ensured their safety.

> *"I had the opportunity to attend many trainings. We went through drills and received training in IPC, Ebola patient management, and other areas. So, I gained both knowledge and confidence in IPC. I already had some understanding of it, but through these trainings, my knowledge improved, and I felt more assured of my own safety while caring for infectious patients."* -Participant 04

### Negative experiences

Five sub-themes emerged from this study; resource constraints, increased workload and stress, psychological and emotional distress, alterations in family and social life, and challenging team dynamics.

The first theme highlighted resource constraints during the Ebola outbreak, encompassing two categories; equipment scarcity and personnel shortage. The second theme focused on increased workload and stress centring on physical strain. The third theme addressed alterations in family and social life featuring two categories; disrupted family life and social isolation. The fourth theme explored the psychological and emotional distress comprising four related categories; fear of infection and death, trauma from witnessing patient death, trauma from severe signs and symptoms of death and post-traumatic stress events. The fifth theme delved into challenging team dynamics, with three categories; compensation and motivation discrepancies, undermining by colleagues, absenteeism from work. Lastly, a related category centring on discomfort wearing personal protective equipment also emerged.

### Resource constraints

#### Equipment scarcity

Participants highlighted significant resource constraints during the early days of the Ebola outbreak. These included a lack of medical equipment, such as personal protective equipment (PPE), which heightened the risk of exposure to the virus. In addition, the facility faced medication shortages and unreliable electricity. The inadequate space and personnel further exacerbated the situation, making the initial period particularly challenging.

> *"You don’t have enough supplies, the space is limited, and there aren’t enough personnel. When you put all that together, especially in the first few days, it makes everything really difficult."* -Participant 06

#### Personnel shortage

Another participant reported that because of the inadequacy of medical equipment, the standard operating procedures were not very stringent at the beginning of the period. Later, however, the Ministry of Health and World Health Organization came in and brought more staff and equipment.

> *"At the beginning, the IPC measures weren’t very stringent. The standard operating procedures were somewhat relaxed because, as you know, when you’re just starting out, not all the necessary human resources or equipment are in place. But as time went on, the Ministry of Health and WHO sent experts to support us in strengthening IPC."* -Participant 04

### Increased workload and stress

#### Physical strain

Many of the participants talked of how they found the first few days of caring for Ebola patients to be extremely exhausting and tiring. They explained that this was because of the emergency nature of the situation which demanded a great deal of effort, leaving them feeling drained and worn out. The rapid setup of units and the need to move suspected cases to isolation wards added to the exhaustion.

> *"My first day in the ETU was exhausting. It all happened as an emergency, and we weren’t prepared for it. We were moving around while the unit was still being set up, and at the same time, we had to transfer suspected cases from the wards to the isolation unit or ETU. It wasn’t an easy situation—it was overwhelming, and there was just too much work."* -Participant 02.

### Psychological and Emotional Distress

#### Fear of infection and death

The fear of contracting and the possibility of dying from EVD, especially after contact with EVD suspects and critically ill patients was a significant source of distress for the participants. Many reported worrying about being the next Ebola patient.

> *"My first day at the ETU was extremely traumatizing because it involved a baby we had already treated in the paediatric ward. Knowing I had previously contact with that child made me very anxious—I kept thinking I was going to get infected too. Treating the same patient again in the ETU was emotionally overwhelming, and I constantly feared I would be the next to test positive."* - Participant 04.

Some participants reported contracting the disease later, which was traumatizing as they feared they would be the next to die, especially after witnessing other health workers succumb to the illness.

> *"Two of our colleagues passed away. At that time, I was already unwell—my result had come back positive. I felt like I was going to be the next to die. It was incredibly traumatizing."* -Participant 02

#### Trauma from witnessing patient death

The nurses witnessed the deaths of many patients, and all reported this as a traumatizing experience. They narrated that the trauma stemmed from the nature of patient deaths with severe symptoms such as bleeding and vomiting and this had a lasting impact on the nurses. Some participants described these days as their worst, and one mentioned that the loss of patients was unlike anything they had ever seen before.

> *"It was so traumatizing seeing people die. They would come in at any stage, often with severe symptoms—vomiting blood, passing blood in their stool. I had never seen someone vomit that much blood before. It was deeply disturbing."* -Participant 02.

> *"The moment I entered, I found people had collapsed—some had already died. It was a mess. There were full buckets of blood, people had passed out stools… it was terrible. I’ve lost patients before, but I had never witnessed anything like that experience."* -Participant 07

In addition, the nurses discussed the unpredictability of the disease, where patients who were expected to survive because they came in stable ended up dying. This added to their trauma.

> *"There was a time when someone came in walking, looking stable, and then kept deteriorating until they died. That experience gave me such a terrible headache—it really affected me."* -Participant 04

#### Trauma from severe signs and symptoms of disease

Several participants reported that the presentation of patients with Ebola virus disease was traumatizing. This presentation included severe vomiting, diarrhoea, and bleeding.

> *"As the number of cases increased and the severity of the disease intensified, patients were bleeding, having diarrhoea, and vomiting constantly. We were overwhelmed, caring for people in that state. By the time you entered the patient’s room, you’d find pools of vomit and blood—but you had to clean them and try to make them feel better. It was truly something disastrous."* - Participant 04

#### Post-traumatic stress events

Some participants reported that the severe symptoms and death that they witnessed while caring for patients with EVD elicited post-traumatic stress events. They further mentioned that did not want to recall or discuss these experiences due to the lasting mental distress.

> *"The things we saw while caring for patients—the blood, the vomit, the diarrhoea, the death—those are things I don’t want to remember. There are some things I can’t even talk about because I never want to see them again… it still disturbs my mind."* -Participant 07.

### Alterations in family and social life

#### Disrupted family life

The outbreak also disrupted participants’ family lives. Some nurses chose not to disclose to their family members that they were working in the ETU to avoid causing fear and anxiety among their family members.

> *"That’s one thing I never did—I never told them I was working in the ETU. I was just surviving on my own. I only said I was in a place where there was Ebola and that I was working at the hospital, like they already knew. But I didn’t want them to be too anxious, so I kept the full truth from them."* - Participant 04

Others who informed their family members about their work reported that their families became very worried about the possibility of contracting and dying from the disease. This caused fear and anxiety among the nurses and affected their productivity. As a result, some nurses stopped communicating with their families when their family members became overly concerned.

> *"At home, everyone was worried… I couldn’t pick up their calls because every time I spoke to them; I felt like crying. So, I chose not to talk to them. They kept calling, but I left my phones busy—I knew that talking to them would only make them worry more. So, I just kept all my feelings to myself."* -Participant 06.

One participant reported that they could barely leave the hospital premises because of the fear that they could spread the disease to others unknowingly.

> *"I stayed alone in the house—I didn’t go to the shop or anywhere else. When you’re working during such an outbreak, you’re constantly thinking that you’re sick."* -Participant 06.

#### Social isolation

Participants also reported being ostracized by people from the community who would run away whenever they came back from duty due to the fear that they could be collected as suspects.

> *"Whenever people saw that bus, they thought it had come to pick up Ebola suspects—so they would run away from me. They didn’t realise I was the one being dropped off. People were really afraid of it."* -Participant 07

### Challenging team dynamics

#### Absenteeism from work

The participants shared negative experiences about working with their colleagues in the ETU. They complained about their colleagues frequently not showing up for work, especially at the beginning of the period when staff numbers were already limited, which made the situation worse.

> *"So, you would find that by the time you entered, others had already left. You were supposed to work with someone, but they weren’t there—so you ended up doing tasks alone, and that really put you at risk."* -Participant 03.

#### Undermining by colleagues

There was undermining from the senior staff. Undermining was especially done by new health staff brought by the Ministry of Health and World Health Organization, most of whom had engaged in previous Ebola outbreaks elsewhere.

> *"I think it wasn’t easy, because when some of the new team members came in, it felt like they were undermining the work we had already done on the ground. They acted as if we knew nothing about IPC—just because they had managed Ebola before, maybe in Sierra Leone or Liberia."* -Participant 06

#### Compensation and motivation discrepancies

Disparities in compensation and working conditions between recruits and existing staff led to feelings of unfairness and demotivation among the participants. It was reported that the recruits were compensated with a better pay than them, the existing staff. In addition, the participants reported that the recruits had better motivation and working conditions.

> *"What I saw as a challenge was that most of the people who came from outside the hospital seemed more motivated than those of us already working here. That didn’t sit well with me, especially since we were doing the same duties. The only difference was that they had come from different areas. But the recruits were paid more—for us, it was 80,000* (Uganda shillings) *per day, while for them, I think it was around 250,000* (Uganda shillings)*. You can see the difference."* -Participant 07.

#### Discomfort wearing personal protective equipment

Wearing PPE was physically uncomfortable and heightened the participants’ anxiety, particularly during the initial days when they were not accustomed to the gear.

> *“My dear, it was a terrible experience, if I may say. I remember putting on those overalls, masks, and goggles—it felt like I was suffocating. I think it was because of the fear, especially since I hadn’t entered the isolation area before. The first time I wore that gear, I was sweating and struggling to breathe properly."* -Participant 07.

## Discussion

Our study engaged nurses who provided care to EVD patients and documented their lived experiences during the Ebola outbreak in Uganda. The findings reveal a range of emotional, psychological, social, and practical challenges these nurses faced, such as resource constraints, increased workload, psychological distress, and complex team dynamics, all of which significantly shaped their frontline experience.

Despite these challenges, positive factors like patient recovery, a supportive work environment, and empowering opportunities played a crucial role in bolstering the resilience and motivation of these nurses.

Nurses caring for EVD patients reported significant physical exhaustion, primarily due to the sudden onset of the outbreak, which caught healthcare facilities unprepared and drastically increased their workload. This is not different from findings from earlier studies on the outbreak of infectious diseases like Ebola (23, 24), indicating that nurses experience extreme physical fatigue and discomfort during disease outbreaks. This was related to the intense work and overwhelming numbers of patients. During such times, nurses have had to assume multiple roles to accommodate the unexpected surges in patients and increased demand for healthcare services (24, 25). However, a contrasting study from Mbarara during the same outbreak found that patient numbers were reduced (12). This difference in findings could be attributed to the fact that there was no Ebola outbreak in Mbarara(7); instead, people feared visiting the hospital due to concerns about isolation and being labelled as EVD suspects(12).

In this study, nurses described being traumatized by the severe signs and symptoms of the disease, as well as by patient deaths. The traumatic impact of patient death has been corroborated by other studies, which have linked this trauma to factors such as the patient’s age, the manner of death, the number of fatalities, and the nurse’s relationship with the patient(26, 27). Additionally, nurses expressed fear of acquiring and dying from EVD, a sentiment echoed in previous research where the uncertainty surrounding EVD heightened this fear(28). Continuously training nurses to improve their knowledge and skills regarding epidemic disease management and increased clinical experience could help mitigate these fears and better equip them to cope with future outbreaks(28).

Reports on the scarcity of medical equipment and personnel highlighted resource constraints. These findings are comparable to those of similar studies in Liberia which reported shortages of personnel protective equipment (PPE), staff, and treatment beds (28, 29). Nurses in this study noted that the lack of PPE forced them to improvise, increasing their risk of EVD exposure. Additionally, the inadequate staffing, compounded by absenteeism, led to longer shifts and the assumption of multiple roles by the remaining staff. This shortage was largely due to healthcare workers’ reluctance to work in ETUs out of fear of infection(24, 30). Such conditions heighten the risk of exposure to infectious body fluids increasing the likelihood of disease transmission among staff (31). These findings underscore the urgent need for adequate resource allocation and staffing during epidemics to safeguard the well-being of frontline nurses and ensure effective patient care(32).

Also, nurses reported that their family lives had been affected, as many had to stay away from their families due to the fear surrounding the disease and its contagious nature. Similar concerns have been documented in previous studies, where nurses expressed worries about the safety of their families due to the threat posed by epidemics(30). Some were even discouraged by their families from working in EVD patient wards(30, 33), while others had to live separately from their families during infectious disease outbreaks(28). This separation often forced nurses to weigh their commitment to work against their responsibilities to their families (34). These challenges led many to experience distress, with some even considering resignation (30). The emotional toll of these experiences highlights the need for robust psychosocial support systems for healthcare workers during epidemics to help them cope with the strain of being separated from their families.

Conflicts between old and new staff are common during epidemics (35). Nurses in this study expressed frustration over the discrepancies in compensation and motivation between new recruits and existing staff. Such disparities can lead to reduced morale and commitment among existing staff(36), potentially impacting the quality of care provided. Moreover, there were reports of undermining behaviour by new health staff, particularly those with previous experience in other outbreaks. Such behaviour not only affects the work environment but also poses risks to patient safety, as it can result in decreased collaboration and communication among healthcare workers(37, 38). This highlights the need for fair and transparent compensation among epidemic response teams to maintain a cohesive and effective workforce.

On a positive note, the nurses shared stories of joy from patient progress, such as improvement, recovery, and discharge. These experiences align with another study in which health workers reported experiencing joy in seeing patients improve and successfully carrying out their duty of caring for them (39). This sense of joy impacts positively on patient care(23), but also positively impacts the nurses’ well- being (24).

A positive work environment is crucial for providing high-quality care, reducing hospital-acquired infection rates, hospital mortality, and adverse events. It is also linked to the recruitment and retention of healthcare professionals during epidemics (40). In this study, the nurses reported experiencing a supportive work environment. They worked together as a team, supporting, watching, and protecting one another.

These findings are consistent with studies from Sierra Leone, where health workers reported a similar positive team environment, with managers providing regular communication, encouragement, and support(28).

Moreover, the nurses in this study felt that the IPC training significantly enhanced their professional knowledge and boosted their confidence in caring for EVD patients. This is consistent with prior studies showing that these trainings help to improve nurses’ skills and knowledge in triage, management of Ebola, and IPC measures(28, 41). This improves service provision for patients, reduces infection spread among nurses, and better equips nurses to manage future outbreaks(41). This has been evidenced by nurses in Sierra Leone and the Democratic Republic of Congo stating that the knowledge and skills gained from IPC training during the Ebola epidemic were utilized to handle the COVID pandemic (42). These findings underscore the importance of continuous professional development in preparing healthcare workers for future epidemics.

From the study findings, the following recommendations were made. The ministry 6of Health should implement fair and transparent distribution of risk allowances to all frontline workers and introduce non-monetary incentives such as medals, certificates of recognition, and public acknowledgment of their service. This will boost their morale and commitment encouraging their participation in future health emergencies. It is crucial to establish a robust rapid response system for infectious disease outbreaks, including the prompt deployment of officials, support staff, and necessary equipment. This is critical for managing and containing diseases before they spread uncontrollably. They should also implement ongoing training programs focusing on epidemic management, particularly for diseases like Ebola. These programs should emphasize infection prevention and control (IPC) measures so that healthcare workers can swiftly implement them in both routine and emergency scenarios.

Nurses are encouraged to engage in continuous education and training to enhance their knowledge and skills in managing epidemic diseases. They should also consistently follow IPC protocols in their everyday patient care. By being diligent in these efforts, they will be better prepared to manage and contain infectious diseases during epidemics, thus ensuring the safety of both patients and healthcare workers.

The study faced several limitations. We acknowledge that our study was constrained by a small sample size, which did not allow for data saturation, a key aspect of qualitative research. Nonetheless, this size was sufficient to identify significant themes related to the research questions. The study specifically targeted nurses who provided care to EVD patients at MRRH, meaning the findings may not reflect the experiences of the other nurses who provided care to EVD patients but weren’t stationed at MRRH. Additionally, the study did not include other health workers involved in the care of EVD patients. They may have had a different experience as they provided care. Despite these limitations, our study made a valuable contribution by examining the real-life experiences of nurses caring for patients with Ebola virus disease, filling an important gap in the literature. Our use of qualitative methods allowed for a thorough exploration of the nurses’ experiences, providing insights that quantitative methods could not capture.

## Conclusion

The study highlights the difficulties experienced by nurses caring for EVD patients. However, it also identifies positive factors that played a crucial role in maintaining their resilience and dedication. The findings emphasize the necessity of a balanced approach to epidemics, one that not only tackles the challenges faced but also reinforces and utilizes the positive factors to improve overall effectiveness and well-being in future health crises. Providing fair compensation, comprehensive psychosocial support, and ongoing training can significantly enhance the resilience, performance and well-being of frontline staff in future outbreaks.

## Data Availability

All data produced in the present study are available upon reasonable request to the authors

## List of abbreviations

CDC: Centres for Disease Control and Prevention
COVID-19: Corona Virus Disease – 2019
ETU: Ebola treatment units
EVD: Ebola Virus Disease
IPC: Infection Prevention and Control
IRB: Institutional Review Board
MRRH: Mubende Regional Referral Hospital
PPE: Personal protective equipment
SSA: Sub-Saharan Africa
SHSREC: School of Health Sciences Research Ethics Committee
WHO: World Health Organization

## Declarations

### Ethics approval and consent to participate

This study protocol and tools were approved by the School of Health Sciences Research and Ethics Committee (SHSREC) of Makerere University (MAKSHSREC- 2023-494). Additionally, permission to conduct the study was granted by the Mubende Hospital Research and Ethics Committee. Each participant gave written informed consent prior to participation. Involvement in the study was voluntary and there were no repercussions for non-participation. All methods were carried out in accordance with the Helsinki declaration and other relevant national guidelines and regulations.

## Consent for publication

Not applicable

## Availability of data and materials

The datasets used and/or analysed during the current study are available from the corresponding author on reasonable request.

## Competing interests

The authors declare that they have no competing interests.

## Funding

This was a self-funded study.

## Authors’ contributions

**GM** Conception and design, acquisition, analysis, interpretation of the data, and manuscript drafting. **SJN** Analysis, interpretation of the data, and manuscript drafting. **RN** Field supervision, data analysis, data interpretation, and critical revision of the manuscript. **BA** Analysis, interpretation of the data, and manuscript drafting. **PM** Supervision, Conception and design, validation of data analysis, data interpretation, and critical revision of the manuscript. **MN** data analysis and data interpretation. All authors accept responsibility for all aspects of the work, including ensuring that any questions about the accuracy or integrity of any part of the work are thoroughly investigated and resolved.

## Acknowledgments

The authors acknowledge the management of the Mubende Regional Referral Hospital for permitting us to conduct this study and the participants who tolerated the long interviews and shared their lived experiences.

## Notes

### Competing Interest Statement

The authors have declared no competing interest.

### Author Declarations

The School of Health Sciences Research and Ethics Committee of Makerere University gave ethical approval for this work (MAKSHSREC-2023-494).

